# The landscape of pediatric procedural sedation in UK & Irish emergency departments; a PERUKI study

**DOI:** 10.1101/2022.06.15.22276454

**Authors:** Dani Hall, Tadgh Moriarty, Damian Roland, Ronan O’Sullivan, Carol Blackburn, Stuart Hartshorn, Shrouk Messahel, Mark D Lyttle

## Abstract

**Study objective:** Approximately 250,000 children undergo pediatric procedural sedation (PPS) in UK & Irish emergency departments (ED) annually. PPS practice in our setting has not been described as fully as in other high income countries. We aimed to evaluate PPS in UK and Irish EDs.

**Methods:** Online survey distributed through Pediatric Emergency Research in the UK and Ireland (PERUKI) during June 2020. One respondent per ED completed the survey, including questions on agents, fasting, training and governance. Results are presented using descriptive statistics.

**Results:** 61/72 (85%) sites responded, of which PPS was performed in 50 (82%). Intravenous ketamine was the most common agent (43/50; 86%), followed by variable concentration nitrous oxide (13/50; 26%). Fasting practices varied widely across sites and agents: 24/45 (53.3%) of sites delivering ketamine/es-ketamine PPS required fasting compared to 2/13 (15.4%) before variable concentration nitrous oxide. 49/72 (68.1%) provided complete responses on training; internal training packages existed in under half (22/49, 44.9%). Most had a guideline (43/61; 70.5%) and documentation proforma (39/61; 63.9%). Databases existed in 24/61 (39.3%).

**Conclusion:** We have demonstrated widespread PPS use, but non-standardized practice. This leads to potential issues of risk and variability, highlighting a need for a UK and Ireland sedation package to standardize PPS practice and data collection, informed by international guidance and evidence. We propose development of a prospective ED sedation registry to facilitate data collection to support research within this area.

## INTRODUCTION

### Background

Many painful and anxiety-provoking procedures in children, such as laceration repair and fracture reduction, can be safely performed in Emergency Departments (ED) with the use of pediatric procedural sedation (PPS), with benefits for patients, families, and health services. These include obviation of the need for general anesthesia, with same-day performance of a definitive procedure avoiding a later return and in-patient hospital stay. This enhances patient care, optimizes convenience for families, and reduces costs and crowding, with up to a tenfold reduction in time to fracture management and ED length of stay compared with general anesthesia.^1^ PPS has been practiced in EDs worldwide for several decades and is included in the curriculum for subspecialty pediatric emergency medicine training internationally.^2–4^ However, despite national guidance, barriers to performing PPS in EDs in the UK and Ireland have been acknowledged, including lack of training and education, and resistance from other specialties.^5^ There is however a growing drive to perform PPS in EDs. For example the recent British Orthopedic Association Standard on pediatric forearm fractures promotes early manipulation and casting without necessitating admission.^6^

### Importance

Although other international groups have described their practice,^7–10^ no published literature exists to evaluate the agents, doses, practices and training programs in place in the UK and Ireland. Paediatric Emergency Research in the UK and Ireland (PERUKI) highlighted PPS as an area of priority for research,^11,12^ and a recently published survey of pain management in the PERUKI network showed that only one-third of sites offer PPS.^13^ Developing a greater understanding of the obstacles and enablers to PPS, practice in regards to agents and procedures, and standards for governance and training are essential to underpin service changes and future research studies.

### Goals of this investigation

In this online survey we therefore aimed to further evaluate practice in PPS in EDs in the UK and Ireland across an international research network. Anticipated findings included identification of areas for improvement in PPS practice and training to enable standardization in line with latest evidence and highlight ways to address barriers in PPS delivery.

## METHODS

### Study design and setting

This online survey study was delivered between 24th June and 23rd July 2020. This was a survey of departmental practice which did not contain patient data, completed by members of a professional network; formal ethical approval exemption was confirmed by the NHS Health Research Authority. Those invited were provided with information regarding the study, and participation was taken as consent. Results are reported in line with the Checklist for Reporting Results of Internet E-Surveys, CHERRIES, statement (appendix 1).^14^ The full survey is available in the online supplemental appendix 2.

### Survey development

Survey content was developed iteratively, using evidence identified from a literature search of international PPS practice. Refinements were made based on input from the study team and PERUKI, and a pilot phase ensured comprehensiveness and wording were appropriate. Information was sought regarding PPS practices, staffing, training, fasting, clinical monitoring, individual sedation agents, and governance and audit activity. The survey included single-select, multiple-select and free-text data collection fields. Adaptive questioning was used to reduce respondent fatigue.

Agents surveyed included ketamine, es-ketamine, midazolam, propofol, ketofol, fixed concentration nitrous oxide, variable concentration nitrous oxide, and intranasal opiate.

Clinicians involved in administering PPS were categorized as consultants (senior doctors who have completed specialty (board level) training), registrars (typically having between 6 and 10 years postgraduate experience, senior house officers (typically three to five years postgraduate experience) and advanced nurse practitioners.

### Survey distribution and participants

Survey responses were collected and managed using Research Electronic Data Capture (REDCap) tools, a secure web-based software platform designed to support data capture for research studies.^15,16^ An individualized electronic link was sent to PPS leads across PERUKI, with one response sought from each site. PERUKI is a research collaborative consisting of sites providing tertiary and secondary level care, in urban and rural settings. In sites where PPS was not practiced, the survey was completed by the PERUKI site lead. A maximum of three reminders were sent to site leads, two weeks and three weeks after opening, and 48 hours before closing. Responses were stored on a secure server in the University of Bristol; whilst participants provided contact information to aid clarification on responses if necessary, data were anonymized at the point of extraction prior to analysis by the study team.

### Data analysis

Data were analyzed using Microsoft Excel, and results are presented using descriptive statistics. Categorical variables are presented using frequency and proportion, and continuous variables are presented using median and ranges where appropriate. Where free text responses were provided, these were analyzed to establish any common themes and coded within the overall data where possible and relevant.

Both partial and complete responses are included. Results pertaining to survey sections are presented with denominators which reflect the number of sites which provided a complete response by section.

## RESULTS

### Demographics

In total 61/72 (84.7%) sites responded. Of these, 47 (77.0%) were mixed adult and pediatric EDs, while 14 (22.9%) were dedicated pediatric EDs; 28 (45.9%) were tertiary hospitals, 32 (52.5%) were secondary hospitals, and one (1.6%) private secondary hospital.

PPS was performed in 50/61 sites (82.0%); 9/61 (14.8%) used PPS ‘daily’, 17 (27.9%) ‘a few times a week’, eight (13.1%) used it ‘weekly’, and one ‘monthly’ or less frequently.

In the five years prior to COVID-19, 35/61 (57.4%) reported an increasing frequency of PPS, while 10 (16.4%) reported a decline. Frequency was unchanged in the remaining sixteen. Of 55 sites who responded to questions regarding the impact of COVID-19 on PPS, one-quarter (13/55, 23.6%) reported a rise in PPS activity, while 23 (41.8%) reported a decrease.

Intravenous ketamine was the most commonly used agent (40/50, 80.0%), followed by variable concentration nitrous oxide (13/50, 26.0%) (**table 1**). Of note, 18/50 sites (36.0%) considered single agent intranasal opiate (diamorphine or fentanyl) a sedative rather than purely an analgesic, while 35/50 (70%) sites considered fixed concentration nitrous oxide a sedative rather than an anxiolytic/analgesic.

**Table 1.** Demographics of sites surveyed, procedural sedation agents and non-pharmacological adjuncts. Key: Trauma Network Designation: MTC, Major Trauma Centre, a designated hospital with all the facilities and specialities to treat patients with any type of injury or combination; MTU, Major Trauma Unit, a designated hospital within a trauma network that provides treatment for most injured patients; Neither, hospitals in the Republic of Ireland where a trauma network is not yet in existence. Pharmacological Agents: VCNO, variable concentration nitrous oxide, delivered via a flow meter up to 70%; IN opiate, intranasal opiate (fentanyl or diamorphine); Ketofol, ketamine and propofol mix.

|  | Number of Sites (n=61) | Percentage |
| --- | --- | --- |
| **DEMOGRAPHICS OF SITES SURVEYED** |  |  |
| **Location** |  |  |
| England | 48 | 78.7% |
| Ireland | 5 | 8.2% |
| Scotland | 5 | 8.2% |
| Wales | 2 | 3.3% |
| Northern Ireland | 1 | 1.6% |
| **Type of hospital** |  |  |
| DGH | 32 | 52.5% |
| Tertiary | 28 | 45.9% |
| Non-publicly funded hospital | 1 | 1.6% |
| **Trauma Network Designation** |  |  |
| TU | 35 | 57.4% |
| MTC | 21 | 34.4% |
| Neither | 5 | 12.2% |
| **Setting** |  |  |
| Mixed with separate PED | 46 | 75.4% |
| Paediatric | 14 | 22.9% |
| Mixed with combined PED | 1 | 1.6% |
| **PROCEDURAL SEDATION AGENT** |  |  |
| **Pharmacological Agents** |  |  |
| Ketamine/Es-Ketamine | 45 | 73.8% |
| Ketamine | 43 | 70.5% |
| VCNO | 13 | 21.3% |
| Midazolam | 10 | 16.4% |
| Propofol | 10 | 16.4% |
| VCNO combined with IN opiate | 4 | 6.6% |
| Es-Ketamine | 2 | 3.3% |
| Ketofol | 0 | 0% |
| **Non-Pharmacological Agents** |  |  |
| Distraction Techniques | 46 | 75.4% |
| Play Specialists | 26 | 42.6% |
| Virtual Reality Headsets | 1 | 1.6% |

### Barriers and enablers

Clinical service pressure was the most frequently reported obstacle to PPS (31/61, 50.8%), while the greatest enabler was the opinion held by ED staff that the ED is an appropriate location for PPS (15/61, 24.6%) (**figure 1**).

**Figure 1.**
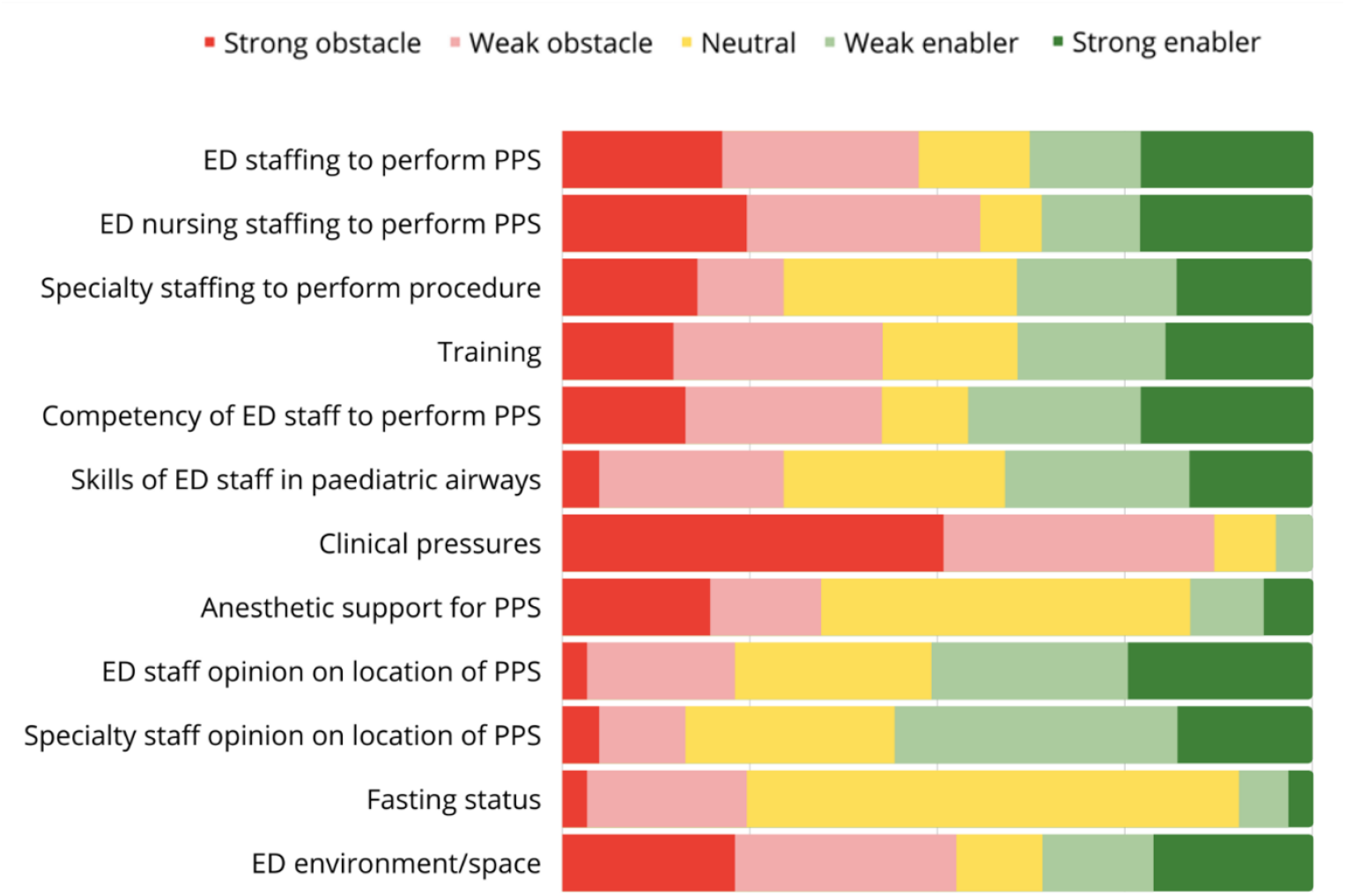
Perceived barriers and enablers to delivery of pediatric procedural sedation in the emergency department, n=61. Key: ED, emergency department; PPS, pediatric procedural sedation

Training was commonly reported as a barrier (26/61, 42.6%), along with ED staff competency (25/61, 41.0%) and pediatric airway management (19/61, 31.1%). Fasting status was considered a barrier in almost a quarter of departments [14/61 (23.0%)].

### Staffing

The use of parenterally administered drugs was associated with the highest proportion of senior clinicians as sedationist (consultant, registrar, or advanced nurse practitioner) with propofol at 100% (10/10), followed by ketamine/es-ketamine at 97.8% (44/45). Inhaled therapies were widely delivered by nurses and non-consultant grade doctors with variable concentration nitrous oxide at 10/13 (76.9%) (**figure 2**).

**Figure 2.**
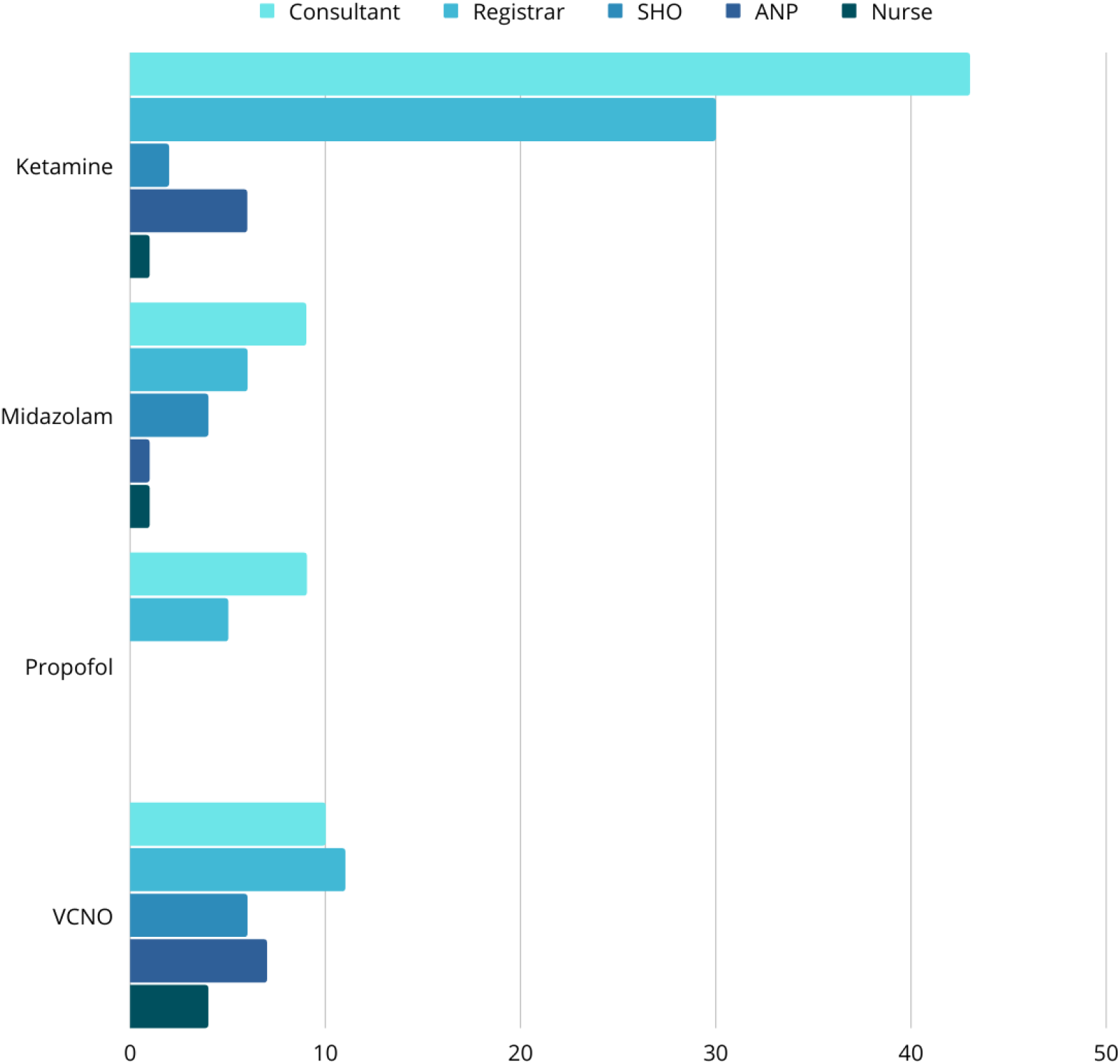
Grade of clinician to take on the role of sedation clinician by agent. Key: Consultant, senior doctor who has completed specialty (board level) training; registrar, typically 6 to 10 years postgraduate experience; SHO, senior house officer, typically 3 to 5 years postgraduate experience; ANP, advanced nurse practitioner.

### Training and governance

Of 49/61 (80.3%) sites providing responses on training, under half had internal training packages (21/49, 42.9%). These included delivering PPS under supervision (21/49, 42.9%), observing a competent provider deliver PPS (16/49, 32.7 %), lectures (7/49, 14.3%), simulation (11/49, 22.4%), a written or online test (7/49, 14.3%) and airway competency sessions in theatre (2/21, 4.1%)

Most sites delivering PPS had a guideline (43/50, 86.0%), checklist for contraindications (36/50, 72.0%), PPS documentation proforma (39/50, 78.0%) and equipment checklist (36/50, 72.0%). Half (26/50, 52.0%) had a clinical practice guideline on management of complications, and most (39/50, 78.0%) did not utilize a PPS-specific sedation adverse event reporting tool.

Discharge criteria were explicit in many sites using parenteral sedation including ketamine / es-ketamine (40/45, 88.9%), midazolam (9/10, 90.0%), propofol (7/10, 70%), and variable concentration nitrous oxide (9/13, 69.2%). Sedation audit databases were maintained in 24/50 (48%) sites, with a minority (4/50, 8.0% sites) collecting qualitative or quantitative data on parental or staff satisfaction.

### Procedures facilitated

The most frequent procedures facilitated were wound closure (31/50, 62.0%), dislocation reduction (28/50, 56.0%), fracture reduction (28/50, 56.0%), foreign body removal (17/50, 34.0%) and CT imaging (12/50, 24.0%) (**table 2**).

**Table 2.** Procedures performed by sedation agent, number (percentage) Key: FCNO, fixed concentration nitrous oxide (50%); VCNO, variable concentration nitrous oxide (max 70%); CT, computerised tomography; MRI, magnetic resonance imaging

| Procedure | Ketamine/<br>Es-ketamine<br>n=45 | FCNO<br>without<br>opiate<br>n=30 | FCNO<br>with<br>opiate<br>n=26 | VCNO<br>n=13 | Midazola<br>m<br>n=10 | Propofol<br>n=10 | Proportio<br>n of sites<br>n=61 |
| --- | --- | --- | --- | --- | --- | --- | --- |
| Wound closure | 30 | 27 | 19 | 10 | 4 | 0 | 46 |
| Nailbed repair | 13 | 13 | 7 | 6 | 1 | 0 | 2 |
| Foreign body removal | 24 | 24 | 8 | 10 | 3 | 0 | 38 |
| Fracture reduction | 43 | 18 | 22 | 8 | 5 | 7 | 48 |
| Dislocation reduction | 42 | 22 | 22 | 9 | 6 | 8 | 50 |
| Dressing change | 9 | 25 | 16 | 11 | 3 | 0 | 38 |
| Lumbar puncture | 1 | 4 | 1 | 4 | 2 | 0 | 8 |
| CT imaging | 8 | 0 | 0 | 0 | 4 | 1 | 10 |
| MRI imaging | 1 | 0 | 0 | 0 | 2 | 1 | 4 |
| Peripheral cannulation | 0 | 9 | 0 | 4 | 3 | 0 | 14 |

### Consent

Most sites required written (33/50, 66.0%) or verbal (40/50, 80.0%) consent, with written consent most commonly obtained for ketamine/es-ketamine (34/45, 75.6%) or propofol (6/10, 60.0%) (**table 3**).

**Table 3.** Doses, age cut-offs, monitoring, consent and location by agent, n (%) unless otherwise stated. Key: Es-ket, es-ketamine; VCNO, variable concentration nitrous oxide; IV, intravenous; IM, intramuscular; IN, intranasal; PO, per oral; NIBP, non-invasive blood pressure; h, hour; m, months; y, years; $, clinician discretion

|  | Ketamine/ Es-ketamine<br>n=45 (74%) |  |  |  |  | VCNO<br>n=13 (21%) | Propofol<br>n=10 (16%) | Midazolam<br>n=10 (16%) |  |  |
| --- | --- | --- | --- | --- | --- | --- | --- | --- | --- | --- |
| Route | Ketamine<br>IV<br>n=40 (66%) | Ketamine<br>IM<br>n=13 (21%) | Ketamine<br>IN<br>n=1 (2%) | Es-ket IV<br>n=2 (7%) | Es-ket IM<br>n=1 (2%) |  | IV | IV<br>n=7<br>(11%) | PO<br>n=6<br>(10%) | Buccal<br>n=6<br>(10%) |
| Initial dose (mg/kg)<br>median (range) | 1<br>(0.5 – 1.5) | 2.5<br>(1 – 5) | 4 | 0.375<br>(0.25 – 0.5) | 3.5<br>(3 – 4) | 50% - 70% | 1<br>(0.5 – 2) | 0.1<br>(0.025–0.1) | 0.5<br>(0.25–0.5) | 0.25<br>(0.1–0.5) |
| Maximum dose<br>(mg/kg)<br>median (range) | 2<br>(1 - 5) | 4<br>(3.5 – 10) | 4 | 3<br>(1-5) | 4 | 70% | 1<br>(0.5-2) |  |  |  |
| Age limitations<br>present<br>median (range) | 43 (96)<br>1y (3m–5y) |  |  |  |  | 11 (85)<br>1y (2m–6y) | 9 (90)<br>12y (2–12y) | 8 (80)<br>1y (6m– 1y) |  |  |
| Consent<br>written<br>verbal<br>none | 33 (73)<br>12 (27)<br>0 (0) |  |  |  |  | 7 (54)<br>5 (38)<br>1 (8) | 6 (60)<br>4 (40)<br>0 | 3 (30)<br>7 (70)<br>0 |  |  |
| Monitoring<br>heart rate<br>NIBP<br>respiratory rate<br>oximetry<br>capnography | 45 (100)<br>38 (84.4)<br>41 (91.1)<br>45 (100)<br>33 (73.3) |  |  |  |  | 13 (100)<br>5 (38.5)<br>9 (69.2)<br>13 (100)<br>3 (23.1) | 10 (100)<br>9 (90.0)<br>10 (100)<br>10 (100)<br>10 (100) | 10 (100)<br>8 (80.0)<br>8 (80.0)<br>10 (100)<br>5 (50.0) |  |  |
| Location<br>resuscitation bay<br>HDU<br>majors cubicle<br>minors area<br>sedation room | 40 (88.9)<br>9 (20.0)<br>4 (8.9)<br>0<br>8 (17.8) |  |  |  |  | 10 (76.9)<br>3 (23.1)<br>3 (23.1)<br>2 (15.4)<br>5 (38.5) | 10 (100)<br>0<br>0<br>0<br>0 | 8 (80.0)<br>2 (20.0)<br>5 (50.0)<br>0<br>3 (30.0) |  |  |

**Table 4.** Fasting requirement by sedation agent, number (percentage) and median (range). Key: h, hours; VCNO, variable concentration nitrous oxide (maximum 70%)

|  | **Ketamine/**<br>**es-ketamine**<br>**n=45 (74%)** | **Propofol**<br>**n=10**<br>**(16%)** | **Midazolam**<br>**n=10**<br>**(16%)** | **VCNO**<br>**n=13**<br>**(21%)** |
| --- | --- | --- | --- | --- |
| **Fasting** |  |  |  |  |
| Always | 8 (17.8%) | 2 (20.0%) | 2 (20.0%) | 1 (7.7%) |
| Sometimes | 16 (35.6%) | 6 (60.0%) | 5 (50.0%) | 1 (7.7%) |
| Never | 21 (46.7%) | 2 (20.0%) | 3 (30.0%) | 11 (84.6%) |
| **Clear fluids** | 2h (1–3h) | 2h | 2h | 2h |
| **Breast milk** | 2h (1–4h) | 2h (2–6h) | 2h (2–4h) | 3h (2–4h) |
| **Infant formula** | 4h (1–6h) | 4h (2–6h) | 4h (2–6h) | 3h (2–6h) |
| **Solids** | 4h (2–6h) | 4h (2–6h) | 6h (2–6h) | 3h (2–6h) |

### Sedation agents

Ketamine/es-ketamine was the most frequently delivered PPS agent in 45/50 (90.0%). A small number of sites adjusted the dose by age (4/45, 8.9%) or following opiate administration (5/45, 11.1%), and one-quarter (12/45, 26.7%) routinely co-administered another drug, most commonly ondansetron (6/45, 13.3%). A clinician responsible for delivering ketamine separately to the clinician performing the procedure was required in all sites.

Of 13 sites who reported variable concentration nitrous oxide use, intranasal opiate was routinely co-administered by 4/13 (30.8%), equally split between fentanyl and diamorphine. Midazolam use was infrequently reported (10/61, 16.4%). A variety of routes were noted, most frequently intravenous (7/10, 70.0%). Most sites delivered midazolam in a resuscitation bay (8/10, 80%), and in most there was a separate sedation clinician to the procedure clinician (6/10, 60.0%). Propofol was also infrequently utilized (10/61, 16.4%), with doses mostly adjusted if pre-procedural opiates were given (7/10, 70.0%). Propofol administration was exclusively administered in a resuscitation bay (10/10, 100%). Further data regarding individual agents is reported in **table 3**.

### Fasting

Fasting practice varied widely across sites and agents: 24/45 (53.3%) required fasting before ketamine/es-ketamine, 2/13 (15.4%) before variable concentration nitrous oxide, 8/10 (80.0%) before propofol, and 7/10 (70.0%) before midazolam.

### Monitoring

There was wide variation in monitored physiology between sites and agents. Heart rate and oximetry were monitored in all sites for ketamine / es-ketamine (45/45), propofol (10/10) and midazolam (10/10) PPS. Least monitored was capnography, being measured in 33/45 (73.3%) of ketamine/es-ketamine sedations, 3/13 (23.1%) variable concentration nitrous oxide and 5/10 (50.0%) of midazolam PPS (**Table 3**).

## DISCUSSION

Our survey of the scope and practice of PPS practice in the UK and Ireland has demonstrated wide PPS use, with 82% of responding sites practicing PPS either a few times a week, or daily. However, we have found variation in training and governance, sedation practice and fasting practice.

### Training and governance

In 2016, McCoy et al described barriers to PPS in the UK and Ireland, identifying three main themes: training and education of ED staff, ED staffing and environment and engagement with the wider hospital.^5^ Similar themes were identified in our survey, with clinical pressures, the availability of ED staffing and limitations in the ED environment posing the greatest barriers. Despite the presence of PPS in the Pediatric Emergency Medicine curriculum,^2^ staff competence in PPS and pediatric airway management remain widely cited barriers to PPS delivery in our study. Lack of standardized training is clearly illustrated in our findings, with under half of sites surveyed having internal training packages.

Variation and paucity in training has also been cited by other regions. Despite the inclusion of PPS as a core competency in Pediatric Emergency Medicine fellowship programs in North America,^17,18^ a quarter of respondents (most of whom were consultant-level physicians) reported learning PPS through self-study in one survey.^19^ Of those with formal PPS training, mode of delivery varied widely including classroom lectures, observation of at least 10 sedations, and online learning. Our findings are comparable to other European sites, with a recent research network survey reporting that half the sites performing PPS required sedation clinicians to have undertaken a pediatric sedation and analgesia course as well as hold advanced life support provider status.^7^ Although not unique internationally, paucity of sedation training across the PERUKI network is a key factor leading to barriers to delivery of PPS in EDs.

We found that of the sites providing PPS, a majority had a guideline (43/50, 86.0%), a pre-procedural checklist (41/50, 82.0%) or standardized documentation proforma (39/50, 78.0%). This is comparable to other European sites; guidelines were available in 74% of European sites and pre-procedural checklists in 51%.^7^ Furthermore, less than half of sites in our survey had a guideline for management of complications (26/51, 42.6%). Our data also illustrates that less than half (24/50, 48%) of sites delivering PPS in our cohort maintain a sedation database for audit purposes. Although 86% of sites delivering PPS in our network have a standardized guideline, this illustrates a deficit of PPS guidelines and standardized proformas in up to 14% of sites utilizing PPS in our network. This combined with a deficit in audit practice leads to potential issues of risk and variability in PPS practice.

### Sedation practice

A 2017 PERUKI survey, exploring structure supporting analgesic practices, described delivery of PPS in 13 (37%) of the 40 PERUKI member sites at the time.^13^ In three years there has been a relative increase of 45%. However, despite wide use of PPS, we have found wide variability in agents and practices.

Unlike data from a Canadian registry of intravenous PPS, in which 11.5% of ketamine PPS episodes were co-delivered with fentanyl,^20^ no sites in our survey combined ketamine with fentanyl. We have found that the use of midazolam is far less ubiquitous than in Europe and Australasia; our survey demonstrated 10/61 (16.4%) of sites delivering PPS used midazolam compared to 100% of respondents of a recent European survey and 68/75 (91%) of respondents in an Australasian survey.^7,9^ Our survey also demonstrated low use of propofol in the UK and Ireland: 10/61 (16.4%) compared to 67% of REPEM sites^7^ and only 3.9% of PPS episodes in a Canadian registry.^20^ Of note, Bhatt’s 2018 study demonstrated propofol sedations were associated with the highest number of serious adverse events at 3.7%.^20^ No PERUKI sites delivered ketofol PPS; ketofol was found to have the third highest number of serious adverse events in the Canadian registry at 2.1%.^20^

A minority of sites (5/43, 11.6%) delivering ketamine PPS also gave ondansetron to minimize vomiting. However, Bhatt (2018) demonstrated that pre-procedural antiemetics are significantly associated with decreased odds of vomiting.^20^ This is likely to be a reflection of the 2020 RCEM ketamine guidance which recommends rescue ondansetron for intractable vomiting post-procedure but does not make reference to its prophylactic use.^21^

Of particular note, we found few sites utilizing capnography during PPS (34/45, 73.3%, of sites delivering ketamine; 3/13, 23.1%, for variable concentration nitrous oxide; and 5/10, 50.0%, for midazolam). In 2014 guidance, the American College of Emergency Physicians stated that *“capnography may be used as an adjunct to pulse oximetry and clinical assessment to detect hypoventilation and apnea earlier than pulse oximetry and/or clinical assessment alone in patients undergoing procedural sedation and analgesia in the ED*.*”*^3^ Recent RCEM national ketamine guidance states that capnography should be included in routine monitoring in children undergoing ketamine sedation,^21^ a change from previous RCEM guidance. Our findings demonstrate significant deviation from national guidance in the use of capnography for PPS, although evidence for increased safety in PPS with the use of capnography is lacking.

### Fasting

Whilst traditional anesthetic practice favors fasting prior to PPS, international registry data demonstrate no association between pre-procedural fasting and emesis, aspiration or other adverse events.^20,22–26^

Fasting was cited as a barrier to PPS in a quarter (15/61, 24.6%) of sites in our study, with highly variable fasting practice between both sites and agents, many of which are utilizing close to American Society of Anesthesiologists preoperative fasting recommendations. However, prolonged fasting in PPS has been associated with adverse outcomes in some patients and leads to periods of extended waiting in the ED, impacting patient flow.^20,27,28^

Contemporary guidance states that PPS should not be delayed in the ED in children who are not fasted.^3,21^ An international consensus statement on fasting before procedural sedation in children and adults published in 2020 recommends a risk-stratification approach to fasting.^27^ In this guideline, children with no risk factors (over the age of 12 months without obesity or severe systemic disease) undergoing elective PPS with an agent other than propofol should aim for a fasting time of two hours for solids and non-breast milk and no fasting time for breast milk or clear fluids. Although this is relevant for elective or day case procedures, PPS performed in ED is, by definition, an urgent or emergency procedure. The consensus statement states that children with mild risk factors requiring PPS for an urgent or emergency procedure should not have their procedure delayed based on fasting time.

However, despite this international guidance, we have demonstrated variation in fasting practice across sites. Despite widespread publication of evidence demonstrating safe PPS practice without fasting, reflected in national and international guidance, a significant proportion of sites continue to fast children. This practice is being reported as a barrier to PPS in the ED. Changing practice in line with latest evidence would immediately facilitate a greater use of PPS and align with time-based targets for flow in EDs.

### The impact of the COVID-19 pandemic

An additional question regarding the impact of the COVID-19 pandemic on PPS in their department was added after the survey had opened, capturing data from 55/72 (76.4%) sites. We were interested in exploring the hypothesis that reduced access to general anesthesia would increase the frequency of PPS in EDs, but this was not found in our data. Frequency of PPS decreased in the majority of sites (23/55, 41.8%), or was unchanged (19/55, 34.5%), with a small proportion reporting a rise (13/55, 23.6%). Potential explanations include the reduction seen in injury presentations seen at the onset of the pandemic;^29^ a concern that airway manipulation during a potential adverse event would convert PPS S to an aerosol generating procedure; concerns that delivery of variable concentration nitrous oxide is via an open circuit and therefore aerosolizing; limiting use of personal protective equipment; competing pressures for space; and staff shortages secondary to sickness or secondments to other clinical areas.

### Limitations

Our study has several limitations. The survey was distributed through the PERUKI network which may have led to a biased representation of PPS practice across the UK and Ireland, favoring practice in sites with a research interest. One respondent per site completed the survey, which may mean the results are a reflection of CPG recommendations and not necessarily of true PPS practice. We tried to mitigate this by requesting that PPS leads, or clinicians with a PPS interest, complete the survey, but by definition these are likely to be senior clinicians, skewing results away from more junior clinicians who may be more likely to actually deliver PPS, leading to sampling bias. Finally, in order to get a complete picture of PPS practice, the survey was long. This may have led to some sites not responding due to time limitations, leading to non-response bias. However, we feel the response rate of 84.7% mitigates this to some extent.

### Conclusion

We have demonstrated wide PPS use, but non-standardized practice in several aspects of PPS, including choice of agent, monitoring and fasting, despite national evidence-based PPS guidelines. We have also demonstrated a paucity of sedation training across the PERUKI network, leading to barriers to delivery of PPS in EDs. As a core competency in the subspeciality Pediatric Emergency Medicine curriculum, there is a need for standardized training, available to multidisciplinary staff, in order to help overcome perceived reported barriers to consistent and sustainable delivery of PPS.

Our study highlights a need for a UK and Ireland sedation package to standardize PPS practice and data collection, informed by international guidance and evidence. We propose development of a prospective ED sedation registry to allow focus on clinical service development and hypothesis generation for future research, enabling international comparisons to be made, leading to evidence-based standardization.

## Supporting information

Supplemental appendix. Full survey

## Data Availability

All data produced in the present study are available upon reasonable request to the authors

---

**Appendix 1.** Checklist for Reporting Results of Internet E-Surveys (CHERRIES) Eysenbach G. Improving the quality of Web surveys: the Checklist for Reporting Results of Internet E-Surveys (CHERRIES). J Med Internet Res 2004;6:e34

| Item category | Checklist Item | Description |
| --- | --- | --- |
| Design | Survey design | The target population was Emergency Departments providing care to children and young people, identified through PERUKI (a collaboration of clinicians and academics from England, Ireland, Northern Ireland, Scotland and Wales). One participant was sought per PERUKI member site, either the Paediatric Procedural Sedation (PPS) lead or the PERUKI site lead. |
| Ethics | Ethics approval | No patient identifiable information was collected – rather the survey collected information on department practices. Formal ethical approval exemption was confirmed by the NHS Research Authority. |
|  | Informed consent process | Participants were provided with information by email and on the welcome page. This included details of survey content, and the time expected for completion (approximately 15 – 20 minutes). Respondents were informed that completion of this survey would be taken as implied consent. |
|  | Data protection | Personal information including respondent name, email address and institution were collected in order to clarify any responses if necessary, and to acknowledge contribution where they agreed to do so. Data were stored securely on University of Bristol servers in line with Data Protection regulations, and not used for any purpose other than this study. Data were pseudo anonymised on download, prior to analysis. |
| Development and pretesting | Development and testing | Survey content was developed iteratively, using evidence identified from a literature search of international PPS practice. Refinements were made based on input from the study team and PERUKI, and a pilot phase ensured comprehensiveness and wording were appropriate. |
| Recruitment process | Open survey versus closed survey | Closed survey |
|  | Advertising the survey | The survey was not advertised; the survey link was sent to site leads via email |
| Survey administration | Web/E-mail | This was a web-based survey, using Research Electronic Data Capture tools (REDCap) |
|  | Context | REDCap is a secure online database for electronic data capture, with functions including survey development and delivery (www.project-redcap.org). |
|  | Mandatory/voluntary | Voluntary |
|  | Incentives | There were no incentives offered for participation |
|  | Time/Date | Between 24th June and 23rd July 2020 |
|  | Randomization of items or questionnaires | No randomisation of items was used, due to the structure of the survey – however participants were given the opportunity to save progress and return in order to minimise survey fatigue |
|  | Adaptive questioning | Adaptive questioning was used throughout the survey to minimize questions asked to be only those relevant based on opening questions. |
|  | Number of Items | Dependent on adaptive questioning – maximum 411 fields |
|  | Number of screens (pages) | Dependent on adaptive questioning – maximum 32 sections |
|  | Completeness check | Questions requiring a response were tagged as mandatory, and respondents were asked to review and provide responses for any such fields prior to submission |
|  | Review step | Respondents were able to review and modify responses prior to submission, using navigation buttons |
| Response rate | Unique site visitor | The audit functionality of REDCap allows logging of visits and responses. This was combined with collection of site names to ensure a maximum of one visit (and response) per site was possible |
|  | View rate | Not applicable – respondents were invited via PERUKI |
|  | Participation rate | 61 responses from 72 invited (85% participation rate) |
|  | Completion rate (Ratio of users who finished the survey/users who agreed to participate) | 59 completed surveys of a total of 61 responses (96.7% completion rate) |
| Preventing multiple entries from the same individual | Cookies used | Other methods, including naming of site and respondent, and REDCap logging tools, were utilized |
|  | IP check | Other methods, including naming of site and respondent, and REDCap logging tools, were utilised |
|  | Log file analysis | Other methods, including naming of site and respondent, and REDCap logging tools, were utilised |
|  | Registration | A login was not used - entry to the survey was via a web link emailed to eligible participants. No duplicate entries were found through the survey question detailing the institution. |
| Analysis | Handling of incomplete questionnaires | Two questionnaires contained incomplete information one on section (training in PPS) but complete information for all other sections. These two questionnaires were excluded from analysis of data on training but included in all other elements of the survey data |
|  | Questionnaires submitted with an atypical timestamp | No respondents were removed from the survey for completing the items too quickly. |
|  | Statistical correction | No weighting scheme was used for the analysis of results. |

