## Supplemental appendix. Full survey for "The landscape of pediatric procedural sedation in UK & Irish emergency departments; a PERUKI study"

### The PoPSiCLe site survey

Paediatric Procedural Sedation - Site Survey

Thank you for being site lead for this survey study which aims to determine current practice in paediatric procedural sedation (PPS) across PERUKI. This will help us describe best practice in training, governance, and quality assurance of PPS, and describe areas for future research.

One person from each site should complete this stage, which will take 15-20 minutes. We do ask you to send some guidelines, audits and training packages during the survey, so it may help to have any relevant documents to hand.

For this survey PPS is defined as "administration of a medication to a child to induce a state, ranging from minimal anxiolysis to a state of deep sedation (but not including general anaesthesia) to facilitate a painful procedure such as laceration repair, foreign body removal or fracture manipulation".

Although fentanyl and diamorphine are not sedative drugs, we are interested in finding out whether and how they are being used as part of a PPS package in your departments - though please don't tell us about them if you use them purely for analgesic purposes.

If something important comes up halfway through, you can leave the survey and return later - just remember to save your return code...

We do ask for your name and contact details so that we can get in touch if we need to clarify your answers, and formally acknowledge your input on any published work resulting from this survey.

There is no consent form to sign, as your completion of this survey will be taken as implied consent. Data will be stored securely on University of Bristol servers in line with current regulations, and will not be used for any purpose other than this study.

---

Your name

__________________________________

---

Your email address

__________________________________

Which site are you answering on behalf of?
(Hint: type a few letters from your site to find it quicker)

◯ Addenbrooke's Hospital
◯ Alder Hey Children's Hospital NHS Foundation Trust
◯ Barking, Havering & Redbridge University Hospitals NHS Trust
◯ Birmingham Children's Hospital
◯ Bon Secours Hospital
◯ Bradford Royal Infirmary
◯ Bristol Royal Hospital for Children
◯ Chelsea and Westminster Hospital
◯ Children's Health Ireland at Crumlin
◯ Children's Health Ireland at Tallaght
◯ Children's Health Ireland at Temple Street
◯ City Hospitals Sunderland NHS Foundation Trust
◯ Cork University Hospital
◯ Countess of Chester NHS Foundation Trust
◯ County Durham & Darlington NHS Foundation Trust
◯ Derriford Hospital
◯ Epsom General Hospital
◯ Evelina London Children's Hospital
◯ Forth Valley Hospital
◯ Frimley Park Hospital
◯ Great North Children's Hospital, Newcastle Upon Tyne
◯ Hull Royal Infirmary
◯ James Cook University Hospital
◯ John Radcliffe Hospital
◯ King's College Hospital
◯ Kingston Hospital
◯ Leeds General Infirmary
◯ Leicester Royal Infirmary
◯ Medway Maritime Hospital
◯ Morriston Hospital
◯ Musgrove Park Hospital
◯ Norfolk & Norwich University Hospital
◯ North Manchester General Hospital
◯ North Middlesex Hospital
◯ Northern Devon Healthcare NHS Trust
◯ Northwick Park Hospital
◯ Nottingham University Hospitals NHS Trust
◯ Ormskirk & District General Hospital
◯ Queen Alexandra Hospital
◯ Queen Elizabeth Hospital, Woolwich
◯ Royal Aberdeen Children's Hospital
◯ Royal Alexandra Children's Hospital
◯ Royal Belfast Hospital for Sick Children
◯ Royal Berkshire NHS Foundation Trust
◯ Royal Bolton Hospital
◯ Royal Derby Hospital
◯ Royal Devon and Exeter Hospital
◯ Royal Free Hospital
◯ Royal Hospital for Children, Glasgow
◯ Royal Hospital for Sick Children, Edinburgh
◯ Royal Manchester Children's Hospital
◯ Royal Preston Hospital
◯ Ipswich Hospital
◯ Royal United Hospital
◯ Royal Wolverhampton NHS Trust
◯ Salisbury NHS Foundation Trust
◯ Sheffield Children's Hospital
◯ Southampton Children's Hospital
◯ St George's Hospital
◯ St Mary's Hospital
◯ The Royal London
◯ University College Hospital, London
◯ University Hospital Crosshouse
◯ University Hospital Lewisham
◯ University Hospital of Wales, Cardiff
◯ Watford General Hospital

◯ Western Sussex Hospitals NHS Trust
◯ Wexham Park Hospital
◯ Whittington Health NHS Trust
◯ Other

---

Please tell us the name of your site

________________________________

---

Is PPS used in your department?
(PPS defined as "administration of a medication to a child to induce a state, ranging from minimal anxiolysis to a state of deep sedation (but not including general anaesthesia) to facilitate a painful procedure such as laceration repair, foreign body removal or fracture manipulation".)

◯ Yes
◯ No

---

Which of the following are used to facilitate PPS in your department?

☐ Ketamine
☐ Es-ketamine
☐ Midazolam
☐ Propofol
☐ Ketofol
☐ Variable % nitrous oxide
☐ 50% nitrous oxide (Entonox) without opiate
☐ Combination of 50% nitrous oxide (Entonox) & opiate
☐ Diamorphine (do not select if used purely as an analgesic or if only used in combination with nitrous oxide)
☐ Fentanyl (do not select if used purely as an analgesic or if only used in combination with nitrous oxide)
☐ Other

---

Please give details of any other agents used in PPS in your ED

________________________________

---

Do you use any non-pharmacological adjuncts to facilitate PPS?

☐ Play specialist
☐ Distraction techniques
☐ Yes, other (please give details below)
☐ No

---

Please give details

________________________________

**How much do you agree with the following statement? "The following have been strong obstacles / enablers in delivering PPS in my ED".**

| | Strong obstacle | Weak obstacle | Neutral | Weak enabler | Strong enabler |
|---|---|---|---|---|---|
| ED staffing to perform PPS | ◯ | ◯ | ◯ | ◯ | ◯ |
| ED nursing staff to support PPS | ◯ | ◯ | ◯ | ◯ | ◯ |
| Specialty staffing to perform procedure | ◯ | ◯ | ◯ | ◯ | ◯ |
| Training | ◯ | ◯ | ◯ | ◯ | ◯ |
| Competence of ED staff in PPS | ◯ | ◯ | ◯ | ◯ | ◯ |
| Skills of ED staff in managing paediatric airways | ◯ | ◯ | ◯ | ◯ | ◯ |
| Clinical pressures | ◯ | ◯ | ◯ | ◯ | ◯ |
| Anaesthetic support for delivery of PPS in ED | ◯ | ◯ | ◯ | ◯ | ◯ |
| Opinion of ED staff on the ED being the appropriate location for PPS / procedure | ◯ | ◯ | ◯ | ◯ | ◯ |
| Opinion of specialty staff on the ED being the appropriate location for PPS / procedure | ◯ | ◯ | ◯ | ◯ | ◯ |
| ED environment / space | ◯ | ◯ | ◯ | ◯ | ◯ |
| Fasting times of children | ◯ | ◯ | ◯ | ◯ | ◯ |
| Other - please give details below | ◯ | ◯ | ◯ | ◯ | ◯ |

Please give details of any other obstacles or enablers to implementing PPS that have been encountered in your ED.

__________________________________

How often is PPS used within your ED?

◯ Daily
◯ A few times a week
◯ Weekly
◯ Monthly
◯ Other

Please give details of the frequency PPS is practiced in your department.

__________________________________

Has COVID-19 changed the frequency at which your department uses PPS?

◯ Increased
◯ Decreased
◯ Stayed the same

Over the 5 year period prior to COVID-19 had the frequency of PPS in your ED:

◯ Increased
◯ Decreased
◯ Stayed the same

How is your department's PPS service delivered?

☐ Planned sedation list
☐ Ad hoc sedations when required
☐ Other - please give details

Please give details of any other ways PPS is delivered in your department.

__________________________________

If you have any comments about any of the areas in this section, please comment below:

#### Guidelines

**Does your ED have any of the following guidelines?**

| | Yes | No |
|---|---|---|
| PPS guideline | ◯ | ◯ |
| Equipment checklist | ◯ | ◯ |
| Patient checklist to ensure there are no contraindications | ◯ | ◯ |
| Guideline on management of complications | ◯ | ◯ |
| Sedation record used for documentation purposes | ◯ | ◯ |

If you have any comments about guidelines for PPS, please comment below:

**What procedures are facilitated by PPS in your department? Choose all that apply.**

**Key:**

**Wound: Wound closure**
**Nail: Nailbed repair**
**FB: Foreign body removal**
**Frac: Fracture reduction**
**Disloc: Dislocation reduction**
**Dress: Dressing change**
**LP: Lumbar puncture**
**CT: CT scan**
**MRI: MRI scan**
**US: Ultrasound**
**Cann: Cannulation**

| | Wound | Nail | FB | Frac | Disloc | Dress | LP | CT | MRI | US | Cann | Other |
|---|---|---|---|---|---|---|---|---|---|---|---|---|
| Ketamine | ☐ | ☐ | ☐ | ☐ | ☐ | ☐ | ☐ | ☐ | ☐ | ☐ | ☐ | ☐ |
| Midazolam | ☐ | ☐ | ☐ | ☐ | ☐ | ☐ | ☐ | ☐ | ☐ | ☐ | ☐ | ☐ |
| Propofol | ☐ | ☐ | ☐ | ☐ | ☐ | ☐ | ☐ | ☐ | ☐ | ☐ | ☐ | ☐ |
| Ketofol | ☐ | ☐ | ☐ | ☐ | ☐ | ☐ | ☐ | ☐ | ☐ | ☐ | ☐ | ☐ |
| Variable % nitrous oxide | ☐ | ☐ | ☐ | ☐ | ☐ | ☐ | ☐ | ☐ | ☐ | ☐ | ☐ | ☐ |
| 50% nitrous oxide (Entonox) | ☐ | ☐ | ☐ | ☐ | ☐ | ☐ | ☐ | ☐ | ☐ | ☐ | ☐ | ☐ |
| Diamorphine | ☐ | ☐ | ☐ | ☐ | ☐ | ☐ | ☐ | ☐ | ☐ | ☐ | ☐ | ☐ |
| Fentanyl | ☐ | ☐ | ☐ | ☐ | ☐ | ☐ | ☐ | ☐ | ☐ | ☐ | ☐ | ☐ |
| Es-ketamine | ☐ | ☐ | ☐ | ☐ | ☐ | ☐ | ☐ | ☐ | ☐ | ☐ | ☐ | ☐ |
| Entonox & opiate combined | ☐ | ☐ | ☐ | ☐ | ☐ | ☐ | ☐ | ☐ | ☐ | ☐ | ☐ | ☐ |

What other procedures are facilitated by PPS?

__________________________________

**Which staff are present during PPS? Choose all that apply.**

**Key and definitions:**

**Clincian both: Clinician responsible for both sedation and procedure**
**Sedationist: Clinician responsible for giving sedation, recognising and managing adverse events with no responsibilities for procedure**
**Sedation assistant: Second clinician responsible for assisting sedationist**
**Proceduralist ED: Clinician responsible for the procedure from ED with no responsibilities for sedation**
**Proceduralist specialty: Proceduralist from specialty team with no responsibility for sedation**

| | Clinician both | Sedationist | Sedation assistant | Proceduralist ED | Proceduralist specialty | Other |
|---|---|---|---|---|---|---|
| Ketamine | ☐ | ☐ | ☐ | ☐ | ☐ | ☐ |
| Midazolam | ☐ | ☐ | ☐ | ☐ | ☐ | ☐ |
| Propofol | ☐ | ☐ | ☐ | ☐ | ☐ | ☐ |
| Ketofol | ☐ | ☐ | ☐ | ☐ | ☐ | ☐ |
| Variable % nitrous oxide | ☐ | ☐ | ☐ | ☐ | ☐ | ☐ |
| 50% nitrous oxide (Entonox) | ☐ | ☐ | ☐ | ☐ | ☐ | ☐ |
| Diamorphine | ☐ | ☐ | ☐ | ☐ | ☐ | ☐ |
| Fentanyl | ☐ | ☐ | ☐ | ☐ | ☐ | ☐ |
| Es-ketamine | ☐ | ☐ | ☐ | ☐ | ☐ | ☐ |
| Entonox & opiate combined | ☐ | ☐ | ☐ | ☐ | ☐ | ☐ |

Which other staff are present during PPS?

__________________________________

If you have any comments about staffing during PPS, please comment below:

**Which grade(s) of clinician can take on the role of sedationist during PPS in your ED?**

| | Consultant (or equivalent) | Registrar (or equivalent) | SHO (or equivalent) | Advanced nurse practitioner | Nurse | Other |
|---|---|---|---|---|---|---|
| Ketamine | ☐ | ☐ | ☐ | ☐ | ☐ | ☐ |
| Midazolam | ☐ | ☐ | ☐ | ☐ | ☐ | ☐ |
| Propofol | ☐ | ☐ | ☐ | ☐ | ☐ | ☐ |
| Ketofol | ☐ | ☐ | ☐ | ☐ | ☐ | ☐ |
| Variable % nitrous oxide | ☐ | ☐ | ☐ | ☐ | ☐ | ☐ |
| 50% nitrous oxide (Entonox) | ☐ | ☐ | ☐ | ☐ | ☐ | ☐ |
| Diamorphine | ☐ | ☐ | ☐ | ☐ | ☐ | ☐ |
| Fentanyl | ☐ | ☐ | ☐ | ☐ | ☐ | ☐ |
| Es-ketamine | ☐ | ☐ | ☐ | ☐ | ☐ | ☐ |
| Entonox & opiate combined | ☐ | ☐ | ☐ | ☐ | ☐ | ☐ |

Which other grade(s) of staff can take on the role of sedationist?

__________________________________

**Does a consultant have to be in the same room as the patient while the sedation episode is occurring?**

| | Yes | No | Sometimes |
|---|---|---|---|
| Ketamine | ○ | ○ | ○ |
| Midazolam | ○ | ○ | ○ |
| Propofol | ○ | ○ | ○ |
| Ketofol | ○ | ○ | ○ |
| Variable % nitrous oxide | ○ | ○ | ○ |
| 50% nitrous oxide (Entonox) | ○ | ○ | ○ |
| Diamorphine | ○ | ○ | ○ |
| Fentanyl | ○ | ○ | ○ |
| Es-ketamine | ○ | ○ | ○ |
| Entonox & opiate combined | ○ | ○ | ○ |

If you have any comments about senior supervision during PPS, please comment below:

#### Training

Does your ED or trust deliver specific competency training in PPS?

◯ Yes
◯ No

What is included in your competency training package?

☐ Lectures
☐ Practical sessions
☐ Simulation scenarios in situ in the environment in which paediatric procedural sedation takes place
☐ Simulation scenarios in a simulation suite
☐ Airway sessions in theatre
☐ Written / on-line test e.g. MCQs
☐ Observing paediatric procedural sedations performed by another clinician - please specify number required
☐ Performing paediatric procedural sedation under the supervision of a 'sedation competent' clinician - please specify number required
☐ Other - please give details

How many does the learner need to observe as part of the competency training package?

__________________________________

How many does the learner need to perform under supervision as part of the competency training package?

__________________________________

What else is included in the competency training package?

__________________________________

Who is this training for? Please choose from the following.

☐ Both doctors and nurses
☐ Doctors only
☐ Nurses only
☐ Other

Who else is this training for?

__________________________________

Is the training package the same for doctors and nurses?

◯ Yes
◯ No

Please give details

__________________________________

---

Does your ED require clinicians involved in PPS to have certification in a nationally recognised paediatric life support course such as APLS?

◯ Yes
◯ No

---

Does your ED require clinicians involved in PPS to have completed training in an intensive care unit such as PICU, NICU, ICU?

◯ Yes
◯ No

---

Are you aware of any external courses in PPS? If yes, please give details

__________________________________

---

If you have any comments about PPS training, please comment below:

**Is consent obtained for PPS, separate to consent obtained for the procedure?**

| | Written consent | Verbal consent | No consent |
|---|---|---|---|
| Ketamine | ☐ | ☐ | ☐ |
| Midazolam | ☐ | ☐ | ☐ |
| Propofol | ☐ | ☐ | ☐ |
| Ketofol | ☐ | ☐ | ☐ |
| Variable % nitrous oxide | ☐ | ☐ | ☐ |
| 50% nitrous oxide (Entonox) | ☐ | ☐ | ☐ |
| Diamorphine | ☐ | ☐ | ☐ |
| Fentanyl | ☐ | ☐ | ☐ |
| Es-ketamine | ☐ | ☐ | ☐ |
| Entonox & opiate combined | ☐ | ☐ | ☐ |

**What monitoring is used during PPS in your ED?**

**Key:**

**O2 sats:  Oxygen saturations**
**HR:  Heart rate**
**BP: Blood pressure**
**RR:  Respiratory rate**
**Capno:  Capnography**
**GCS:  GCS**
**AVPU:  AVPU**

| | O2 sats | HR | BP | RR | Capno | GCS | AVPU | Other | None |
|---|---|---|---|---|---|---|---|---|---|
| Ketamine | ☐ | ☐ | ☐ | ☐ | ☐ | ☐ | ☐ | ☐ | ☐ |
| Midazolam | ☐ | ☐ | ☐ | ☐ | ☐ | ☐ | ☐ | ☐ | ☐ |
| Propofol | ☐ | ☐ | ☐ | ☐ | ☐ | ☐ | ☐ | ☐ | ☐ |
| Ketofol | ☐ | ☐ | ☐ | ☐ | ☐ | ☐ | ☐ | ☐ | ☐ |
| Variable % nitrous oxide | ☐ | ☐ | ☐ | ☐ | ☐ | ☐ | ☐ | ☐ | ☐ |
| 50% nitrous oxide (Entonox) | ☐ | ☐ | ☐ | ☐ | ☐ | ☐ | ☐ | ☐ | ☐ |
| Diamorphine | ☐ | ☐ | ☐ | ☐ | ☐ | ☐ | ☐ | ☐ | ☐ |
| Fentanyl | ☐ | ☐ | ☐ | ☐ | ☐ | ☐ | ☐ | ☐ | ☐ |
| Es-ketamine | ☐ | ☐ | ☐ | ☐ | ☐ | ☐ | ☐ | ☐ | ☐ |
| Entonox & opiate combined | ☐ | ☐ | ☐ | ☐ | ☐ | ☐ | ☐ | ☐ | ☐ |

Please give details of any other monitoring used during PPS

__________________________________

If you measure depth of sedation please give details of the sedation scale you use

__________________________________

#### Where in the ED does PPS take place?

| | Sedation / procedure room | Resuscitation room | High dependency area | Majors area | Minors area | Other |
|---|---|---|---|---|---|---|
| Ketamine | ☐ | ☐ | ☐ | ☐ | ☐ | ☐ |
| Midazolam | ☐ | ☐ | ☐ | ☐ | ☐ | ☐ |
| Propofol | ☐ | ☐ | ☐ | ☐ | ☐ | ☐ |
| Ketofol | ☐ | ☐ | ☐ | ☐ | ☐ | ☐ |
| Variable % nitrous oxide | ☐ | ☐ | ☐ | ☐ | ☐ | ☐ |
| 50% nitrous oxide (Entonox) | ☐ | ☐ | ☐ | ☐ | ☐ | ☐ |
| Diamorphine | ☐ | ☐ | ☐ | ☐ | ☐ | ☐ |
| Fentanyl | ☐ | ☐ | ☐ | ☐ | ☐ | ☐ |
| Es-ketamine | ☐ | ☐ | ☐ | ☐ | ☐ | ☐ |
| Entonox & opiate combined | ☐ | ☐ | ☐ | ☐ | ☐ | ☐ |

Please give details of other location(s) PPS occurs in your ED

__________________________________

#### Ketamine

Is there a lower age limit in place for the use of ketamine for PPS? Please give details

__________________________________

By which route is ketamine given to facilitate PPS in your ED?

☐ Intravenous
☐ Intramuscular
☐ Intranasal
☐ Other

Please give details of other routes by which ketamine PPS is given

__________________________________

Are any medications given routinely with ketamine to facilitate PPS in your ED? Please do not include analgesics given prior to sedation for analgesic purposes rather than sedation purposes.

☐ Atropine
☐ Ondansetron
☐ Midazolam
☐ Other drug(s) - please give details below
☐ No

Please give details of any other drugs routinely given with ketamine to facilitate PPS

__________________________________

Are children fasted before given ketamine for PPS?

◯ Yes always
◯ Yes sometimes
◯ No

Please give details

__________________________________

What fasting time is required after solids?

◯ 2 hours
◯ 4 hours
◯ 6 hours
◯ other

Please give details

__________________________________

What fasting time is required after formula milk?

◯ 2 hours
◯ 4 hours
◯ 6 hours
◯ other

Please give details

__________________________________

What fasting time is required after breast milk?

◯ 2 hours
◯ 4 hours
◯ 6 hours
◯ other

Please give details

__________________________________

What fasting time is required after clear fluids?

◯ 2 hours
◯ 4 hours
◯ 6 hours
◯ other

Please give details

__________________________________

How frequently are observations measured during  sedation with ketamine?

◯ 5 minutely
◯ 10 minutely
◯ 15 minutely
◯ other

Please give details

__________________________________

How frequently are observations measured after sedation with ketamine, during recovery, before the child returns to their normal baseline?

◯ 5 minutely
◯ 10 minutely
◯ 15 minutely
◯ other

Please give details

__________________________________

If you have any comments about ketamine PPS in your department, please comment below:

#### Intravenous ketamine

What initial weight per kg dose is used for intravenous ketamine for PPS?

__________________________________

Can subsequent doses be given?

◯ Yes
◯ No

Please specify subsequent doses

__________________________________

Is there a maximum recommended dose of intravenous ketamine for PPS in your ED?

◯ Yes
◯ No

Please specify maximum dose

__________________________________

Are different weight per kg doses used for different age groups of children?

◯ Yes
◯ No

Please give details

__________________________________

Is the dose of intravenous ketamine adjusted if opiate analgesia is given before PPS?

◯ Yes
◯ No

Please give details

__________________________________

If you have any comments about intravenous ketamine PPS in your department, please comment below:

#### Intramuscular ketamine

What initial weight per kg dose is used for intramuscular ketamine for PPS?

__________________________________

Can subsequent doses be given?

- ◯ Yes
- ◯ No

Please specify subsequent doses

__________________________________

Is there a maximum recommended dose of intramuscular ketamine for PPS in your ED?

- ◯ Yes
- ◯ No

Please specify maximum dose.

__________________________________

Are different weight per kg doses used for different age groups of children?

- ◯ Yes
- ◯ No

Please give details

__________________________________

Is the dose of intramuscular ketamine adjusted if opiate analgesia is given before PPS?

- ◯ Yes
- ◯ No

Please give details

__________________________________

If you have any comments about intramuscular ketamine PPS in your department, please comment below:

#### Midazolam

Is there a lower age limit in place for the use of midazolam for PPS? Please give details

__________________________________

By which route is midazolam given to facilitate PPS in your ED?

☐ Intravenous
☐ Oral
☐ Buccal
☐ Intranasal
☐ Other

Please give details of other routes by which midazolam PPS is given

__________________________________

Are any medications given routinely with midazolam to facilitate PPS in your ED? Please do not include analgesics given prior to sedation for analgesic purposes rather than sedation purposes.

◯ Yes
◯ No

Please give details of any other medications routinely given with midazolam to facilitate PPS

__________________________________

Are children fasted before given midazolam for PPS?

◯ Yes always
◯ Yes sometimes
◯ No

Please give details

__________________________________

What fasting time is required after solids?

◯ 2 hours
◯ 4 hours
◯ 6 hours
◯ other

Please give details

__________________________________

What fasting time is required after formula milk?

◯ 2 hours
◯ 4 hours
◯ 6 hours
◯ other

Please give details

__________________________________

What fasting time is required after breast milk?

◯ 2 hours
◯ 4 hours
◯ 6 hours
◯ other

Please give details

__________________________________

What fasting time is required after clear fluids?

◯ 2 hours
◯ 4 hours
◯ 6 hours
◯ other

Please give details

__________________________________

How frequently are observations measured during sedation with midazolam?

◯ 5 minutely
◯ 10 minutely
◯ 15 minutely
◯ other

Please give details

__________________________________

How frequently are observations measured after sedation with midazolam, during recover, before full return to the child's normal baseliney?

◯ 5 minutely
◯ 10 minutely
◯ 15 minutely
◯ other

Please give details

__________________________________

If you have any comments about midazolam PPS in your department, please comment below:

#### Intravenous midazolam

What initial weight per kg dose is used for intravenous midazolam for PPS?

__________________________________

Can subsequent doses be given?

◯ Yes
◯ No

Please specify subsequent doses.

__________________________________

Is there a maximum recommended dose of intravenous midazolam for PPS in your ED?

◯ Yes
◯ No

Please specify maximum dose.

__________________________________

Are different weight per kg doses used for different age groups of children?

◯ Yes
◯ No

Please give details

__________________________________

Is the dose of intravenous midazolam adjusted if opiate analgesia is given before PPS?

◯ Yes
◯ No

Please give details

__________________________________

If you have any comments about intravenous midazolam PPS in your department, please comment below:

#### Oral midazolam

What initial weight per kg dose is used for oral midazolam for PPS?

__________________________________

Can subsequent doses be given?

◯ Yes
◯ No

Please specify subsequent doses.

__________________________________

Is there a maximum recommended dose of oral midazolam for PPS in your ED?

◯ Yes
◯ No

Please specify maximum dose.

__________________________________

Are different weight per kg doses used for different age groups of children?

◯ Yes
◯ No

Please give details

__________________________________

Is the dose of oral midazolam adjusted if opiate analgesia is given before PPS?

◯ Yes
◯ No

Please give details

__________________________________

If you have any comments about oral midazolam PPS in your department, please comment below:

#### Buccal midazolam

What initial weight per kg dose is used for buccal midazolam for PPS?

__________________________________

Can subsequent doses be given?

◯ Yes
◯ No

Please specify subsequent doses.

__________________________________

Is there a maximum recommended dose of buccal midazolam for PPS in your ED?

◯ Yes
◯ No

Please specify maximum dose.

__________________________________

Are different weight per kg doses used for different age groups of children?

◯ Yes
◯ No

Please give details

__________________________________

Is the dose of buccal midazolam adjusted if opiate analgesia is given before PPS?

◯ Yes
◯ No

Please give details

__________________________________

If you have any comments about buccal midazolam PPS in your department, please comment below:

#### Intranasal midazolam

What initial weight per kg dose is used for intranasal midazolam for PPS?

__________________________________

Can subsequent doses be given?

◯ Yes
◯ No

Please specify subsequent doses.

__________________________________

Is there a maximum recommended dose of intranasal midazolam for PPS in your ED?

◯ Yes
◯ No

Please specify maximum dose

__________________________________

Are different weight per kg doses used for different age groups of children?

◯ Yes
◯ No

Please give details

__________________________________

Is the dose of intranasal midazolam adjusted if opiate analgesia is given before PPS?

◯ Yes
◯ No

Please give details

__________________________________

If you have any comments about intranasal midazolam PPS in your department, please comment below:

#### Variable concentration nitrous oxide

Is there a lower age limit in place for the use of variable concentration nitrous oxide for PPS? Please give details

__________________________________

Are children fasted before given variable concentration nitrous oxide for PPS?

◯ Yes always
◯ Yes sometimes - please give details
◯ No

Please give details

__________________________________

What fasting time is required after solids?

◯ 2 hours
◯ 4 hours
◯ 6 hours
◯ other

Please give details

__________________________________

What fasting time is required after formula milk?

◯ 2 hours
◯ 4 hours
◯ 6 hours
◯ other

Please give details

__________________________________

What fasting time is required after breast milk?

◯ 2 hours
◯ 4 hours
◯ 6 hours
◯ other

Please give details

__________________________________

What fasting time is required after clear fluids?

◯ 2 hours
◯ 4 hours
◯ 6 hours
◯ other

Please give details

__________________________________

Are any medications given routinely with variable concentration nitrous oxide to facilitate PPS in your ED? Please do not include analgesics given prior to sedation for analgesic purposes rather than sedation purposes.

◯ Yes
◯ No

Please give details of any medications routinely given with variable concentration nitrous oxide PPS

__________________________________

How frequently are observations measured during sedation with variable concentration nitrous oxide?

◯ 5 minutely
◯ 10 minutely
◯ 15 minutely
◯ other

Please give details

__________________________________

How frequently are observations measured after sedation with variable concentration nitrous oxide, during recovery, before full return to the child's normal baseline?

◯ 5 minutely
◯ 10 minutely
◯ 15 minutely
◯ other

Please give details

__________________________________

If you have any comments about variable concentration nitrous oxide PPS in your department, please comment below:

#### 50% concentration nitrous oxide (Entonox) without opiate

Are children fasted before given Entonox for PPS?

◯ Yes always
◯ Yes sometimes - please give details
◯ No

Please give details

__________________________________

Are any medications given routinely with Entonox to facilitate PPS in your ED? Please do not include opiates in this section.

◯ Yes
◯ No

Please give details of any medications routinely given with Entonox to facilitate PPS

__________________________________

How frequently are observations measured during sedation with Entonox?

◯ 5 minutely
◯ 10 minutely
◯ 15 minutely
◯ other

Please give details

__________________________________

How frequently are observations measured after sedation with Entonox, during recovery?

◯ 5 minutely
◯ 10 minutely
◯ 15 minutely
◯ other

Please give details

__________________________________

If you have any comments about Entonox PPS in your department, please comment below:

#### Propofol

Is there a lower age limit in place for the use of propofol for PPS? Please give details

__________________________________

Are any medications given routinely with propofol to facilitate PPS in your ED? Please do not include analgesics given prior to sedation for analgesic purposes rather than sedation purposes.

◯ Yes
◯ No

Please give details of any medications routinely given with propofol to facilitate PPS

__________________________________

Are children fasted before given propofol for PPS?

◯ Yes always
◯ Yes sometimes - please give details
◯ No

Please give details

__________________________________

What fasting time is required after solids?

◯ 2 hours
◯ 4 hours
◯ 6 hours
◯ other

Please give details

__________________________________

What fasting time is required after formula milk?

◯ 2 hours
◯ 4 hours
◯ 6 hours
◯ other

Please give details

__________________________________

What fasting time is required after breast milk?

◯ 2 hours
◯ 4 hours
◯ 6 hours
◯ other

Please give details

__________________________________

What fasting time is required after clear fluids?

◯ 2 hours
◯ 4 hours
◯ 6 hours
◯ other

Please give details

__________________________________

What initial weight per kg dose is used for propofol for PPS?

__________________________________

Can subsequent doses be given?

◯ Yes
◯ No

Please specify subsequent doses

__________________________________

Is there a maximum recommended dose of propofol for PPS in your ED?

◯ Yes
◯ No

Please specify maximum dose

__________________________________

Is the dose of propofol adjusted if opiate analgesia is given before PPS?

◯ Yes
◯ No

Please give details

__________________________________

How frequently are observations measured during sedation with propofol?

◯ 5 minutely
◯ 10 minutely
◯ 15 minutely
◯ other

Please give details

__________________________________

How frequently are observations measured after sedation with propofol, during recovery, before full return to the child's normal baseline?

◯ 5 minutely
◯ 10 minutely
◯ 15 minutely
◯ other

Please give details

__________________________________

If you have any comments about propofol PPS in your department, please comment below:

#### Ketofol

Is there a lower age limit in place for the use of ketofol for PPS? Please give details

__________________________________

Are any medications given routinely with ketofol to facilitate PPS in your ED? Please do not include analgesics given prior to sedation for analgesic purposes rather than sedation purposes.

◯ Yes
◯ No

Please give details of any medications routinely given with ketofol to facilitate PPS

__________________________________

Are children fasted before given ketofol for PPS?

◯ Yes always
◯ Yes sometimes - please give details
◯ No

Please give details

__________________________________

What fasting time is required after solids?

◯ 2 hours
◯ 4 hours
◯ 6 hours
◯ other

Please give details

__________________________________

What fasting time is required after formula milk?

◯ 2 hours
◯ 4 hours
◯ 6 hours
◯ other

Please give details

__________________________________

What fasting time is required after breast milk?

◯ 2 hours
◯ 4 hours
◯ 6 hours
◯ other

Please give details

__________________________________

What fasting time is required after clear fluids?

◯ 2 hours
◯ 4 hours
◯ 6 hours
◯ other

Please give details

__________________________________

What initial weight per kg dose is used for ketofol for PPS?

__________________________________

Can subsequent doses be given?

◯ Yes
◯ No

Please specify subsequent doses

__________________________________

Is there a maximum recommended dose of ketofol for PPS in your ED?

◯ Yes
◯ No

Please specify maximum dose

__________________________________

Is the dose of ketofol adjusted if opiate analgesia is given before PPS?

◯ Yes
◯ No

Please give details

__________________________________

How frequently are observations measured during sedation with ketofol?

◯ 5 minutely
◯ 10 minutely
◯ 15 minutely
◯ other

Please give details

__________________________________

How frequently are observations measured after sedation with ketofol, during recovery, before full return to the child's baseline?

◯ 5 minutely
◯ 10 minutely
◯ 15 minutely
◯ other

Please give details

__________________________________

If you have any comments about ketofol PPS in your department, please comment below:

#### Es-ketamine

Is there a lower age limit in place for the use of es-ketamine for PPS? Please give details

__________________________________

By which route is es-ketamine given to facilitate PPS in your ED?

☐ Intravenous
☐ Intramuscular
☐ Intranasal
☐ Other

Please give details of other routes by which es-ketamine PPS is given

__________________________________

Are any medications given routinely with es-ketamine to facilitate PPS in your ED? Please do not include analgesics given prior to sedation for analgesic purposes rather than sedation purposes.

☐ Atropine
☐ Ondansetron
☐ Midazolam
☐ Other drug(s) - please give details below
☐ No

Please give details of any other drugs routinely given with es-ketamine to facilitate PPS

__________________________________

Are children fasted before given es-ketamine for PPS?

◯ Yes always
◯ Yes sometimes
◯ No

Please give details

__________________________________

What fasting time is required after solids?

◯ 2 hours
◯ 4 hours
◯ 6 hours
◯ other

Please give details

__________________________________

What fasting time is required after formula milk?

◯ 2 hours
◯ 4 hours
◯ 6 hours
◯ other

Please give details

__________________________________

What fasting time is required after clear fluids?

◯ 2 hours
◯ 4 hours
◯ 6 hours
◯ other

Please give details

__________________________________

How frequently are observations measured during sedation with es-ketamine?

◯ 5 minutely
◯ 10 minutely
◯ 15 minutely
◯ other

Please give details

__________________________________

How frequently are observations measured after sedation with es-ketamine, during recovery, before the child returns to their normal baseline?

◯ 5 minutely
◯ 10 minutely
◯ 15 minutely
◯ other

Please give details

__________________________________

If you have any comments about es-ketamine PPS in your department, please comment below:

#### Intravenous es-ketamine

What initial weight per kg dose is used for intravenous es-ketamine for PPS?

__________________________________

Can subsequent doses be given?

◯ Yes
◯ No

Please specify subsequent doses

__________________________________

Is there a maximum recommended dose of intravenous es-ketamine for PPS in your ED?

◯ Yes
◯ No

Please specify maximum dose

__________________________________

Are different weight per kg doses used for different age groups of children?

◯ Yes
◯ No

Please give details

__________________________________

Is the dose of intravenous es-ketamine adjusted if opiate analgesia is given before PPS?

◯ Yes
◯ No

Please give details

__________________________________

If you have any comments about intravenous es-ketamine PPS in your department, please comment below:

#### Intramuscular es-ketamine

What initial weight per kg dose is used for intramuscular es-ketamine for PPS?

__________________________________

Can subsequent doses be given?

◯ Yes
◯ No

Please specify subsequent doses

__________________________________

Is there a maximum recommended dose of intramuscular es-ketamine for PPS in your ED?

◯ Yes
◯ No

Please specify maximum dose.

__________________________________

Are different weight per kg doses used for different age groups of children?

◯ Yes
◯ No

Please give details

__________________________________

Is the dose of intramuscular es-ketamine adjusted if opiate analgesia is given before PPS?

◯ Yes
◯ No

Please give details

__________________________________

If you have any comments about intramuscular es-ketamine PPS in your department, please comment below:

#### Entonox & opiate combined

Is there a lower age limit in place for the use of Entonox and opiate combined for PPS? Please give details

__________________________________

Are children fasted before given Entonox & opiate combination for PPS?

◯ Yes always
◯ Yes sometimes - please give details
◯ No

Please give details

__________________________________

What fasting time is required after solids?

◯ 2 hours
◯ 4 hours
◯ 6 hours
◯ other

Please give details

__________________________________

What fasting time is required after formula milk?

◯ 2 hours
◯ 4 hours
◯ 6 hours
◯ other

Please give details

__________________________________

What fasting time is required after breast milk?

◯ 2 hours
◯ 4 hours
◯ 6 hours
◯ other

Please give details

__________________________________

What fasting time is required after clear fluids?

◯ 2 hours
◯ 4 hours
◯ 6 hours
◯ other

Please give details

__________________________________

How frequently are observations measured during sedation with Entonox & opiate combined?

◯ 5 minutely
◯ 10 minutely
◯ 15 minutely
◯ other

Please give details

__________________________________

How frequently are observations measured after sedation with Entonox & opiate combined, during recovery, before full return to the child's normal baseline?

◯ 5 minutely
◯ 10 minutely
◯ 15 minutely
◯ other

Please give details

__________________________________

If you have any comments about Entonox & opiate combined PPS in your department, please comment below:

#### Recovery and discharge

**Which staff members recover a child as PPS wears off?**

| | 1:1 doctor | 1:1 nurse | 1:1 doctor and nurse | Nurse caring for more than 1 child | Other |
|---|---|---|---|---|---|
| Ketamine | ○ | ○ | ○ | ○ | ○ |
| Midazolam | ○ | ○ | ○ | ○ | ○ |
| Variable % nitrous oxide | ○ | ○ | ○ | ○ | ○ |
| 50% nitous oxide (Entonox) | ○ | ○ | ○ | ○ | ○ |
| Propofol | ○ | ○ | ○ | ○ | ○ |
| Ketofol | ○ | ○ | ○ | ○ | ○ |
| Diamorphine | ○ | ○ | ○ | ○ | ○ |
| Fentanyl | ○ | ○ | ○ | ○ | ○ |
| Es-ketamine | ○ | ○ | ○ | ○ | ○ |
| Entonox & opiate combined | ○ | ○ | ○ | ○ | ○ |

Please give details of any other staff members who recover a child as PPS wears off

__________________________________

**Are children admitted to an inpatient ward or short stay / observation / clinical decision unit after PPS to allow time for sedation to wear off or side-effects such as vomiting to settle (rather than to allow specialty review post procedure or for other non-PPS related reason)?**

| | Yes, routinely | Yes, if near 4 hour breach | Yes, other reason | No |
|---|---|---|---|---|
| Ketamine | ☐ | ☐ | ☐ | ☐ |
| Midazolam | ☐ | ☐ | ☐ | ☐ |
| Variable % nitrous oxide | ☐ | ☐ | ☐ | ☐ |
| 50% nitrous oxide (Entonox) | ☐ | ☐ | ☐ | ☐ |
| Propofol | ☐ | ☐ | ☐ | ☐ |
| Ketofol | ☐ | ☐ | ☐ | ☐ |
| Diamorphine | ☐ | ☐ | ☐ | ☐ |
| Fentanyl | ☐ | ☐ | ☐ | ☐ |
| Es-ketamine | ☐ | ☐ | ☐ | ☐ |
| Entonox & opiate combined | ☐ | ☐ | ☐ | ☐ |

Please give details of other reasons children are admitted to an inpatient ward or short stay / observation / clinical decision unit after PPS

__________________________________

**Are there set criteria to be met prior to discharge after PPS?**

| | Yes | No |
|---|---|---|
| Ketamine | ◯ | ◯ |
| Midazolam | ◯ | ◯ |
| Variable % nitrous oxide | ◯ | ◯ |
| 50% nitrous oxide | ◯ | ◯ |
| Propofol | ◯ | ◯ |
| Ketofol | ◯ | ◯ |
| Diamorphine | ◯ | ◯ |
| Fentanyl | ◯ | ◯ |
| Es-ketamine | ◯ | ◯ |
| Entonox & opiate combined | ◯ | ◯ |

Please give details.

If you have any comments about admission or discharge following PPS in your department, please comment below:

#### Data collection - governance and cost analysis

Do you use a sedation adverse event reporting tool other than local incident reporting tools to document adverse events during PPS?

◯ Yes
◯ No

Please give details of the adverse event reporting tool used in your department.

__________________________________

Does your department keep a sedation log or database to audit PPS practice?

◯ Yes
◯ No

How are you charging for sedation in your ED?

◯ ED attendance
◯ Locally agreed tariff
◯ Day case
◯ Inpatient admission
◯ Not applicable (Republic of Ireland)
◯ Other

Please give details of how you are billing for PPS in your ED

__________________________________

Has your department or trust collected data on the impact PPS has had in your department on any of the following?

☐ Admissions
☐ Bed days
☐ Theatre spaces
☐ Cost savings for the trust
☐ Yes - other (please give details below)
☐ No

Please give details of any other data your department or trust has collected

__________________________________

Has your department collected any qualitative or quantitative data on parental or staff satisfaction of PPS in the ED?

◯ Yes
◯ No

Please give details of any qualitative or quantitative data your ED has collected on parental or staff satisfaction of PPS

__________________________________

If you have any comments about data collection of PPS in your department, please comment below:

#### Closing page

If you have any other comments about this survey or PPS at your site please add your comments below:
